# Increased AGE-RAGE axis stress in methamphetamine (MA) abuse and MA-induced psychosis: associations with oxidative stress and increased atherogenicity

**DOI:** 10.1101/2023.01.21.23284873

**Authors:** Hussein Kadhem Al-Hakeim, Mazin Fadhil Altufaili, Amer Fadhil Alhaideri, Abbas F. Almulla, Shatha Rouf Moustafa, Michael Maes

## Abstract

**Background and aims:** Methamphetamine (MA)-induced psychosis (MIP) is associated with increased oxidative toxicity (especially lipid peroxidation) and lowered antioxidant defenses. Advanced glycation end products (AGEs) cause oxidative stress upon ligand binding to AGE receptors (RAGE). There are no data on whether MA use may cause AGE-RAGE stress, and whether the latter is associated with MIP.

**Methods:** This case-control study recruited 60 patients with MA use disorder and 30 normal controls and measured serum levels of oxidative stress toxicity (OSTOX, lipid peroxidation), antioxidant defenses (ANTIOX), magnesium, copper, atherogenicity, and AGE, soluble RAGE (sRAGE), and computed a composite reflecting AGE-RAGE axis activity.

**Findings:** MA dependence and use were accompanied by increased AGE, sRAGE, AGE-RAGE, OSTOX/ANTIOX, Castelli risk index 1 and atherogenic index of plasma, indicating that MA causes AGE-RAGE axis stress, oxidative damage, and atherogenicity. The severity of dependence and MA dose were strongly correlated with increased sRAGE concentrations. Increased AGE-RAGE stress was strongly associated with OSTOX, OSTOX/ANTIOX, and MA-induced intoxication symptoms, psychosis, hostility, excitation, and formal thought disorders. We found that 54.8% of the variance in MIP symptoms was explained by the regression on AGE-RAGE, the OSTOX/ANTIOX ratio, lowered magnesium, and increased copper, and that these biomarkers mediated the effects of increasing MA doses on MIP symptoms. We found that 36.0% of the variance in the atherogenicity indices was explained by OSTOX/ANTIOX, AGE-RAGE, and lowered magnesium.

**Conclusions:** MA use causes intertwined increases in AGE-RAGE axis stress and oxidative damage, which together predict the severity of MIP symptoms and increased atherogenicity.

## Introduction

Methamphetamine (MA) abuse is a worldwide public health issue linked to a rise in overdose mortality (1) or death from the consequences of MA intake (2). MA is the most widely abused substance in the world after cannabis (3, 4). Several studies have shown that up to 50% of MA patients may develop MA-induced psychosis (MIP) (5, 6). MIP risk and symptom severity have also been reported to rise with MA use indices, which include drug doses and frequency, dependence features, increased quantity of MA use, and abrupt use of large quantities of the drug (7-11).

MIP and schizophrenia have long been recognized to share clinical features, and therefore MIP is often used as a model of schizophrenia (12, 13). Nevertheless, the precise mechanisms that may explain the onset of MIP are not known. A major theory is that MIP is related to MA-induced neurotoxic effects including through alterations in neurotransmitter signaling, inflammatory pathways, neuroinflammation, neuronal apoptosis, death pathways, mitochondrial dysfunction, endoplasmic reticulum stress, astrocytosis, neurodegenerative processes of cortical interneurons (14-19). One major factor that may drive these neurotoxic effects is oxidative damage due to increased production of reactive oxygen species (ROS) and lowered antioxidant defenses (11). Increased oxidative stress toxicity (OSTOX) was esablished in people with MA substance use disorder (SUD) (20, 21), and MIP (11). The latter authors reported that MIP is predicted by an increased OSTOX/antioxidant (ANTIOX) ratio, determined by a) increased oxidized high-density lipoprotein (oxHDL), oxidized low-DL (oxLDL), malondialdehyde (MDA) and myeloperoxidase (MPO), and b) lowered levels of total antioxidant potential (TAC), glutathione peroxidase (Gpx), catalase, zinc, and HDL cholesterol (HDLc) (11).

Advanced glycation end products (AGEs) are potentially harmful heterogeneous molecules of irreversible products derived from nonenzymatic glycation of the reactive carbonyl group of reducing sugar and nucleic acids, lipids, or proteins (22). The pathological implications of AGEs are ascribed to their ability to produce ROS and nitrogen (RNS) species, as well as oxidative stress and inflammation, leading to structural and functional protein alterations, cellular dysfunction and apoptosis, and ultimately tissue injuries (23). Cross-links formed by the interactions of AGEs with their cell surface receptors for advanced glycation end products (RAGEs) have been found during the development and progression of various aging-related diseases, such as diabetes, cardiovascular complications, kidney malfunctions, osteoporosis, cancer, neurodegenerative diseases, and liver disorders (24, 25). Upon ligand binding, RAGE-mediated intracellular signaling (26) can lead to an increase in the production of ROS and inflammatory cytokines (27, 28). The soluble form of RAGE (sRAGE) is involved in host defenses against infections, inflammation, cardiometabolic illnesses, and age-related diseases (27, 29). AGE-RAGE signalling also plays a role in atherogenity and as a risk factor of coronary artery disease (30, 31). The OSTOX profile established in MA use and MIP, including increased oxLDL and oxHDL levels, coupled with lowered HDLc levels, indicates increased atherogeniticy (11). MA use may cause cardiovascular disease including atherosclerotic plaque formation (32). Moreover, MA abuse may cause changes in calcium, magesium and copper metabolism (33-35), which are associated with schizophrenia (36, 37) and increased atherogenicity (11, 38). However, no previous papers examined AGE, sRAGE, calcium, magnesium, and copper levels in MA dependence and MIP, and their association with OSTOX, ANTIOX, and atherogenicity markers.

Hence, the present study was performed to examine AGE, sRAGE, calcium, magnesium and copper levels in MA SUD and MIP, and their associations with OSTOX, ANTIOX, and atherogenicity as assessed with the Castelli risk index 1 and the atherogenic index of plasma (AIP).

## Material and Methods

### Participants

Sixty patients with MA use disorder participated in the present research, which took place between April 2022 and August 2022 at the Psychiatry Unit of Al-Hussein Medical City in the Kerbala Governorate, Iraq. The patients were diagnosed according to the Diagnostic and Statistical Manual of Mental Disorders (the 5th edition) (DSM-5) as moderate to severe MA substance use disorder (SUD) (39) and the International Classification of Disease as MA SUD (F15.20). The patients were divided depending on the quantity of MA use into two groups: high-MA SUD group (31 patients who took 2.435±0.616 gm of MA/day) and low-MA SUD group (29 patients who took 0.948±0.155 gm of MA/day). Because of the religious climate in Karbala city, we were only able to recruit male addicts. All SUD patients had been abusing MA for at least three months, and an urine MA test was positive. Thirty healthy acquaintances of staff members or patients acted as controls. None of the controls had a current or prior axis I DSM-IV-TR diagnosis or a family history of schizophrenia or psychosis, and none of them had used any psychoactive substances (other than nicotine dependence). SUD patients and controls were not included if they had ever been treated with immunosuppressive drugs or glucocorticoids or if they had been diagnosed with a neurodegenerative or neuroinflammatory disease or condition such as Alzheimer’s disease, Parkinson’s disease, multiple sclerosis, or stroke. Patients with (auto)immune disoders including inflammatory bowel disease, rheumatoid arthritis, chronic obstructive pulmonary disease, psoriasis, or diabetes mellitus were also excluded. We excluded any subject with serum TG higher than 400mg/dl (4.156) to apply the fredwald’s equation (40).

The research adhered to Iraqi and international privacy and ethical regulations. Before participating in this study, all participants and first-degree relatives of participants with MA SUD (legal representatives are mother, father, brother, spouse, or son) gave written informed consent. The study was approved by the institutional review board (IRB) of the College of Science, University of Kufa, Iraq (89/2022), Karbala Health Directorate-Training and Human Development Center (Document No.18378/ 2021), which follows the Declaration of Helsinki’s International Guideline for Human Research Protection.

### Clinical assessments

To collect patient and control data, a senior psychiatrist with expertise in SUD conducted a semi-structured interview including rating scales. The severity of the Dependence Scale (SDS) was used to estimate the severity of MA dependence (41). As previously explained, we were able to extract one validated and replicable principal componenet (PC) from 4 SDS items, namely SDS1 (did you ever think your use of MA was out of control), SDS2 (did the prospect of missing MA make you very anxious or worried), SDS4 (did you wish you could stop) and SDS5 (how difficult would you find it to stop or go without MA) and labeled this construct “PC_SDS” (11). Age at onset, duration of MA dependence, daily dosage (gr), route of administration (no=0, orally ingested=1, smoked or snorted=2, and injected=3) were also recorded. As explained previously, we were able to extract one validated and replicable PC from PC_SDS, MA dosage, MA use last month, and route, dubbed PC_MA (11).

### Biomarkers assays

Fasting venous blood was drawn from all participants shortly after admission into hospital. Blood was allowed to coagulate for 10 minutes at room temperature before being centrifuged at 3000 rpm for 10 minutes. The serum was separated and transported to Eppendorf tubes, where it was kept at -80 °C until analysis. The urine MA test was performed immediately by the Multi-Drug 12 Drugs Rapid Test Panel (Urine) kit supplied by Citest Diagnostics Inc., Vancouver, Canada. Serum levels of AGE, sRAGE, catalase, GPx, MPO, malondialdehyde (MDA), OxHDL, OxLDL, and TAC, were measured using commercial ELISA kits supplied by Nanjing Pars Biochem Co., Ltd., Nanjing, China. All kits were based on a sandwich technique and showed an inter-assay CV of less than 10%. Zinc levels were tested spectrophotometrically in serum using an Agappe Diagnostics^®^ ready-to-use kit from Cham (Switzerland). Copper was determined spectrophotometrically using a kit from LTA Co. (Milano, Italy). Total cholesterol, triglycerides (TGs), glucose, and high-density lipoprotein cholesterol (HDLc) were measured using a kit supplied by Spinreact®, Gerona, Spain. Serum calcium and magnesium concentrations were determined using spectrophotometric kits provided by Biolabo^®^ (Maizy, France). The intra-assay coefficients of variation (CV) for all the assays were <10.0% (precision within-assay).

As explained previously (11), we used the following composite scores: a) as an integrated index of AGE-RAGE axis stress, we computed a z composite score as z AGE + z sRAGE, labeled AGE-RAGE axis (ARA) stress, b) OSTOX computed as z MPO + z oxHDL + z oxLDL + z MDA; c) ANTIOX as z catalase + z Gpx + z TAC + z zinc + z HDLc (11); d) the OSTOX/ANTIOX ratio as z OSTOX – z ANTIOX (11), e) as a composite of AGE-RAGE stress + oxidative stress we computed z(AGE+sRAGE) + zOSTOX (labeled: AROSTOX); and f) as a composite of AGE+RAGE stress + oxidative stress / antioxidants ratio we computed z(AGE+sRAGE) + zOSTOX -zANTIOX (labeled: AROSTOX/ANTIOX). The Castelli"s Risk Index-I was calculated as z TC – z HDLc (labeled as Castelli), and AIP as z TG – HDLc (labelled as AIP) (42, 43).

### Statistical analysis

Analysis of variance (ANOVA) was employed to examine differences between groups in continuous variables and analysis of contingency tables (χ^2^-test) to investigate the association between nominal variables. Furthermore, Pearson’s product-moment correlation coefficients were employed to examine the correlation between the two variables. We utilized a multivariate general linear model (GLM) while controlling for confounders variables, such as age, sex, and TUD, to delineate the association between the study group (healthy control/lowMA/highMA), and biomarkers. Multiple regression analysis (manual method) has been utilized to examine whether the biomarkers can significantly predict psychotic symptoms. In addition, we also used a stepwise automated approach with a p-value of 0.05 for entry and 0.1 for removal from the model. Standardized beta coefficients with t statistics and exact p-value were computed for each of the predictors’ variables, and for the model we computed F, statistics and total variance explained (R^2^), which was used as effect size. Furthermore, we consistently used the variance inflation factor and tolerance to examine collinearity and multicollinearity issues. We tested for heteroskedasticity using the White and modified Breusch-Pagan homoscedasticity tests. All statistical test were performed at p=0.05 (two tailed) using IBM Windows SPSS version 28. A priori estimation of the sample size (for multiple regression analysis with up to 6 predictors, alpha=0.05, two tailed, effect sice=0.15, and power=0.8) shows that the sample size should be at least n=75 (G*Power 3.1.9.4).

Using Partial Least Squares (PLS) analysis, the causal associations between MA dose and the severity of the psychotic phenome of MIP was examined, whereby AGE, sRAGE, ARA, OSTOX, ANTIOX, OSTOX/ANTIOX, magnesium, calcium, copper and atherogenicity were considered as mediators of the effects of MA dose on the phenome. All variables were entered as single indicators except the phenome, which was conceptualized as a latent vector extracted from psychosis, hostility, excitement, mennerism (PHEM), formal thought disorders and MAI symptoms, and and atherogenicity which was a LV extracted from AIP and Castelli. When the following conditions are met, we conduct a full PLS analysis: a) adequate construct and convergence validity as indicated by average variance extracted (AVE) > 0.5, rho A > 0.8, Cronbach’s alpha > 0.7, and composite reliability > 0.7, b) all loadings on the latent vectors must be greater than 0.6 at p < 0.001, c) construct’s cross-validated redundancy must be sufficient as indicated by blindfolding analysis, d) the results of the Confirmatory Tetrad Analysis show that the model is not misspecified as a reflective model; and e) the model fit is adequate with standardized root squared residual (SRMR) < 0.08. After confirming the model’s quality based on the aforementioned criteria, we performed a complete PLS-SEM pathway analysis (SmartPLS) using 5,000 bootstraps to calculate the path coefficients (with p-values), specific and total indirect (mediated) effects, and total effects.

## Results

### Sociodemographic data

**Table 1** shows the demographical and clinical parameters in MA SUD patients divided into those with high-MA and low-MA use and healthy controls. The high-MA abuse group took 2.435±0.616 gm/day in comparison with 0.948±0.155 gm/day taken by the low-MA abuse group. No significant differences in age, BMI, marital status, residency, lifetime cannabis use, age at onset of MA use were found among the three study groups. The high-MA abuse group showed a significantly greater unemployment rate as compared with controls and the low-MA group. The duration of MA use (in months) was significantly longer in the high-MA abuse group than in the low-MA abuse group. The route of MA administration is significantly different between patients groups with higher injection intake in the high-MA abuse group. The total SDS score, PC_SDS, and PC_MA abuse, MAI index, psychosis, hostility, excitement, formal thought disorders, OSTOX, and OSTOX/ANTIOX were significantly different among the three study groups and increased from controls → low-MA → high MA abuse. In contrast, the ANTIOX composite decreased from controls → low-MA abuse group → high-MA abuse group.

**Table 1.** Demographics and clinical data in healthy controls (HC) and patients with methamphetamine (MA) abuse and dependence divided into those with high- and low-MA abuse (median split)

| **Variables** | **HC**<br>**(n=30)<sup>A</sup>** | **Low-MA abuse**<br>**(n=29)<sup>B</sup>** | **High-MA abuse**<br>**(n=31)<sup>C</sup>** | **F/$\chi^2$** | **df** | **p** |
| --- | --- | --- | --- | --- | --- | --- |
| MA dose (gm/day) | 0<sup>B, C</sup> | 0.948±0.155<sup>A, C</sup> | 2.435±0.616<sup>A, B</sup> | KWA |  | <0.001 |
| Age (years) | 27.3±5.4 | 25.1±6.6 | 27.9±5.9 | 1.815 | 2/87 | 0.169 |
| BMI (Kg/m<sup>2</sup>) | 25.30±2.81 | 25.40±3.92 | 24.81±3.29 | 0.287 | 2/87 | 0.751 |
| Marital state (Married/Single) | 8/22 | 12/17 | 9/22 | 1.682 | 2 | 0.413 |
| Residency (Urban/Rural) | 23/7 | 20/9 | 20/11 | 1.952 | 2 | 0.329 |
| Employment (N/Y) | 8/22<sup>C</sup> | 8/21<sup>C</sup> | 27/4<sup>A, B</sup> | 29.306 | 2 | <0.001 |
| TUD (N/Y) | 15/15<sup>B, C</sup> | 7/22<sup>A, C</sup> | 1/30<sup>B, C</sup> | 17.578 | 2 | <0.001 |
| Alcohol intake (N/Y) | 30/0<sup>B, C</sup> | 21/8<sup>A, C</sup> | 7/24<sup>B, C</sup> | 41.066 | 2 | <0.001 |
| Duration of MA use (months) | - | 15.03±14.571<sup>C</sup> | 34.03±17.352<sup>B</sup> | 20.941 | 1/88 | <0.001 |
| Age of onset MA abuse (yrs) | - | 23.93±5.861 | 25.19±5.192 | 0.782 | 1/88 | 0.380 |
| MA administration route (Mo/Sm/In) | - | 6/14/7<sup>C</sup> | 11/7/12<sup>B</sup> | 48.151 | 6 | <0.001 |
| Lifetime cannabis use (N/Y) | 0/30 | 0/29 | 3/28 | 5.907 | 2 | 0.104 |
| Total SDS score | 0<sup>B, C</sup> | 9.550±1.298<sup>A, C</sup> | 10.130±1.335<sup>A, B</sup> | KWA |  | <0.001 |
| PC_SDS | -1.334±0<sup>B, C</sup> | 0.593±0.284<sup>A, C</sup> | 0.736±0.241<sup>A, B</sup> | KWA |  | <0.001 |
| PC_MA abuse | -1.382±0<sup>B, C</sup> | 0.548±0.170<sup>A, C</sup> | 0.825±0.192<sup>A, B</sup> | KWA |  | <0.001 |
| MAI symptom index | -1.059±0<sup>B, C</sup> | 0.039±0.751<sup>A, C</sup> | 0.988±0.555<sup>A, B</sup> | KWA |  | <0.001 |
| Psychosis | -1.170±0<sup>B, C</sup> | 0.286±0.739<sup>A, C</sup> | 0.864±0.487<sup>A, B</sup> | KWA |  | <0.001 |
| Excitement | -1.038±0<sup>B, C</sup> | 0.335±0.803<sup>A, C</sup> | 0.692±0.827<sup>A, B</sup> | KWA |  | <0.001 |
| Hostility | -1.102±0.091<sup>B, C</sup> | 0.351±0.775<sup>A, C</sup> | 0.738±0.709<sup>A, B</sup> | KWA |  | <0.001 |
| Formal Thought Disorders | -1.138±0<sup>B, C</sup> | 0.151±0.669<sup>A, C</sup> | 0.960±0.530<sup>A, B</sup> | KWA |  | <0.001 |
| OSTOX | -0.709±0.628<sup>B, C</sup> | 0.096±0.962<sup>A, C</sup> | 0.596±0.918<sup>A, B</sup> | 18.299 | 2/87 | <0.001 |
| ANTIOX | 0.652±0.593<sup>B, C</sup> | -0.056±0.872<sup>A, C</sup> | -0.579±1.069<sup>A, B</sup> | 15.376 | 2/87 | <0.001 |
| OSTOX/ANTIOX | -0.889±0.459<sup>B, C</sup> | 0.099±0.730<sup>A, C</sup> | 0.767±0.926<sup>A, B</sup> | 39.333 | 2/87 | <0.001 |

BMI: body mass index, AGE: advanced glycation end products, AIP: atherogenic index of plasma, ANTIOX: index of antioxidant defenses, KWA: Kruskal-Wallis ANOVA, OSTOX: index of oxidative stress, MAI: MA intoxication symptoms, RAGE: receptor of advanced glycation end products, PC\_MA: MA use severity index, PC\_SDS: index of severity of dependence, SDS: severity of dependence scale, TUD: tobacco use disorder.

### Biomarkers in controls and MA SUD patients

The levels of serum biomarkers in controls and both patient groups are presented in **Table 2**. There is a significant increase in the levels of the following parameters with the highest value in the high-MA group to the lowest values in controls: AGEs, sRAGE, ARA, AROSTOX, and AROSTOX/ANTIOX. HDLc was significantly lower in the high-MA group than in controls. The low-MA group showed significantly higher fasting blood glucose levels than the control group. Both patient groups have significantly higher copper, total cholesterol, triglycerides, VLDLc, Castelli and AIP levels than controls. Both MA groups showed significantly lower magnesium than controls.

**Table 2.**
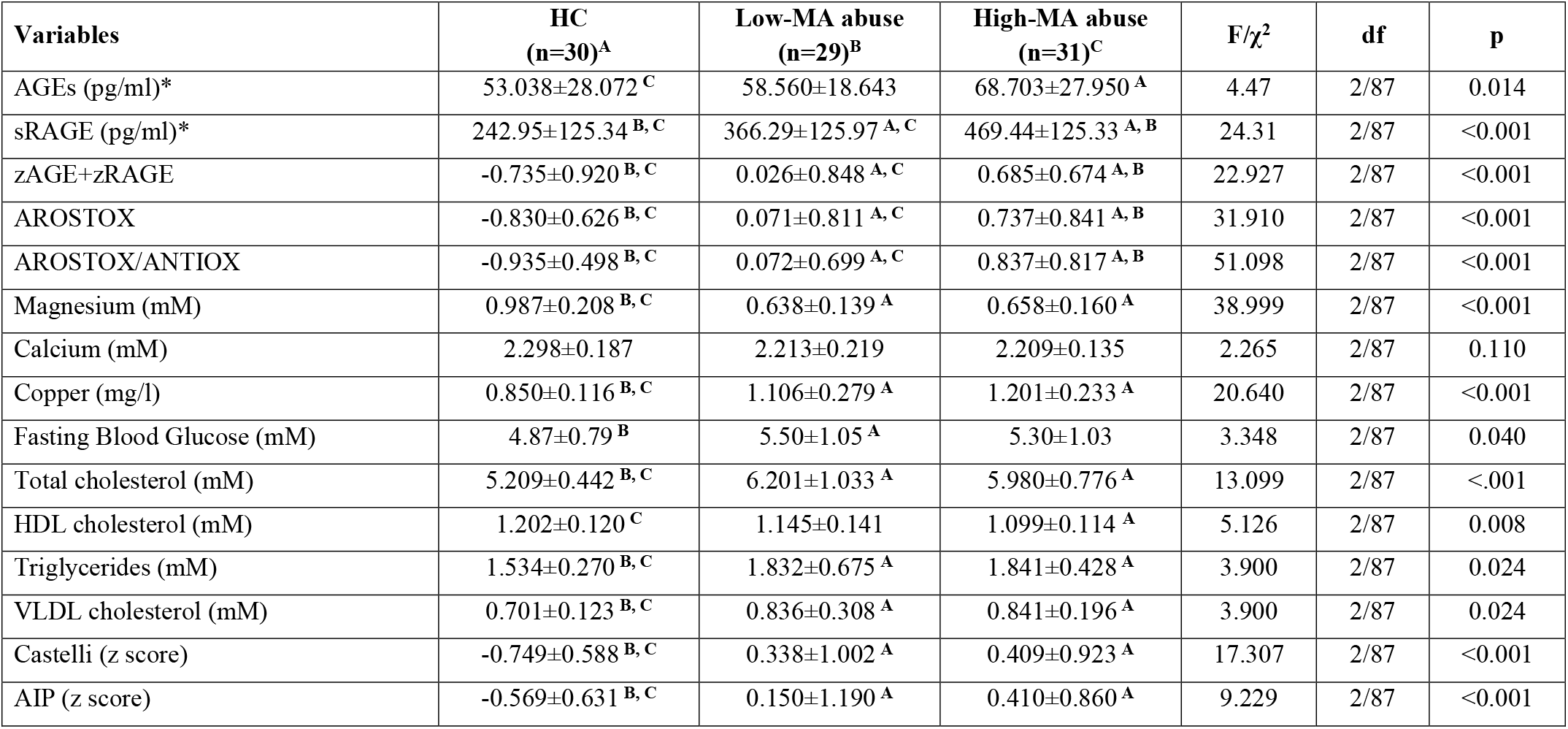
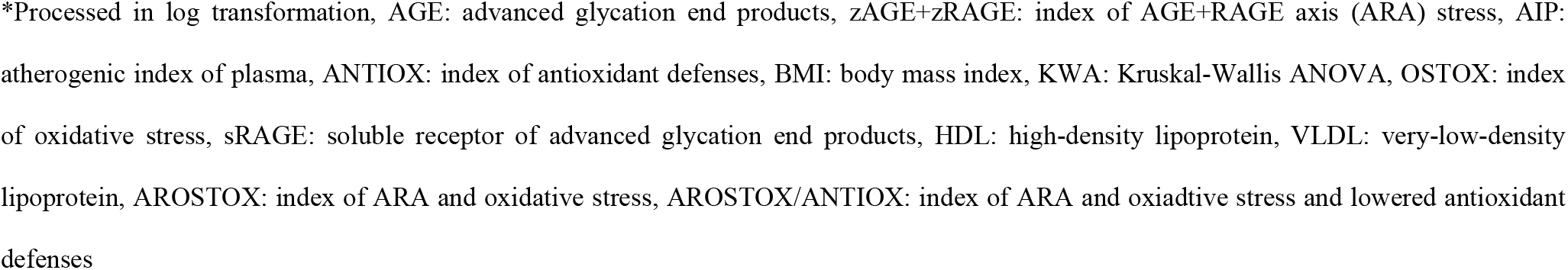
Serum biomarkers in healthy controls (HC) and patients with methamphetamine (MA) abuse or dependence divided into those with high- and low-MA abuse (median split)

| **Variables** | **HC**<br>**(n=30)<sup>A</sup>** | **Low-MA abuse**<br>**(n=29)<sup>B</sup>** | **High-MA abuse**<br>**(n=31)<sup>C</sup>** | **F/$\chi^2$** | **df** | **p** |
| --- | --- | --- | --- | --- | --- | --- |
| AGEs (pg/ml)* | 53.038±28.072 <sup>C</sup> | 58.560±18.643 | 68.703±27.950 <sup>A</sup> | 4.47 | 2/87 | 0.014 |
| sRAGE (pg/ml)* | 242.95±125.34 <sup>B, C</sup> | 366.29±125.97 <sup>A, C</sup> | 469.44±125.33 <sup>A, B</sup> | 24.31 | 2/87 | <0.001 |
| zAGE+zRAGE | -0.735±0.920 <sup>B, C</sup> | 0.026±0.848 <sup>A, C</sup> | 0.685±0.674 <sup>A, B</sup> | 22.927 | 2/87 | <0.001 |
| AROSTOX | -0.830±0.626 <sup>B, C</sup> | 0.071±0.811 <sup>A, C</sup> | 0.737±0.841 <sup>A, B</sup> | 31.910 | 2/87 | <0.001 |
| AROSTOX/ANTIOX | -0.935±0.498 <sup>B, C</sup> | 0.072±0.699 <sup>A, C</sup> | 0.837±0.817 <sup>A, B</sup> | 51.098 | 2/87 | <0.001 |
| Magnesium (mM) | 0.987±0.208 <sup>B, C</sup> | 0.638±0.139 <sup>A</sup> | 0.658±0.160 <sup>A</sup> | 38.999 | 2/87 | <0.001 |
| Calcium (mM) | 2.298±0.187 | 2.213±0.219 | 2.209±0.135 | 2.265 | 2/87 | 0.110 |
| Copper (mg/l) | 0.850±0.116 <sup>B, C</sup> | 1.106±0.279 <sup>A</sup> | 1.201±0.233 <sup>A</sup> | 20.640 | 2/87 | <0.001 |
| Fasting Blood Glucose (mM) | 4.87±0.79 <sup>B</sup> | 5.50±1.05 <sup>A</sup> | 5.30±1.03 | 3.348 | 2/87 | 0.040 |
| Total cholesterol (mM) | 5.209±0.442 <sup>B, C</sup> | 6.201±1.033 <sup>A</sup> | 5.980±0.776 <sup>A</sup> | 13.099 | 2/87 | <.001 |
| HDL cholesterol (mM) | 1.202±0.120 <sup>C</sup> | 1.145±0.141 | 1.099±0.114 <sup>A</sup> | 5.126 | 2/87 | 0.008 |
| Triglycerides (mM) | 1.534±0.270 <sup>B, C</sup> | 1.832±0.675 <sup>A</sup> | 1.841±0.428 <sup>A</sup> | 3.900 | 2/87 | 0.024 |
| VLDL cholesterol (mM) | 0.701±0.123 <sup>B, C</sup> | 0.836±0.308 <sup>A</sup> | 0.841±0.196 <sup>A</sup> | 3.900 | 2/87 | 0.024 |
| Castelli (z score) | -0.749±0.588 <sup>B, C</sup> | 0.338±1.002 <sup>A</sup> | 0.409±0.923 <sup>A</sup> | 17.307 | 2/87 | <0.001 |
| AIP (z score) | -0.569±0.631 <sup>B, C</sup> | 0.150±1.190 <sup>A</sup> | 0.410±0.860 <sup>A</sup> | 9.229 | 2/87 | <0.001 |

\*Processed in log transformation, AGE: advanced glycation end products, zAGE+zRAGE: index of AGE+RAGE axis (ARA) stress, AIP: atherogenic index of plasma, ANTIOX: index of antioxidant defenses, BMI: body mass index, KWA: Kruskal-Wallis ANOVA, OSTOX: index of oxidative stress, sRAGE: soluble receptor of advanced glycation end products, HDL: high-density lipoprotein, VLDL: very-low-density lipoprotein, AROSTOX: index of ARA and oxidative stress, AROSTOX/ANTIOX: index of ARA and oxidative stress and lowered antioxidant defenses

### Results of multivariate GLM analysis

**Table 3** presents the results of multivariate GLM analyses examining the associations between AGE, sRAGE and OSTOX and diagnosis (healthy control / low-MA / high-MA). Multivariate GLM analysis showed that there were significant associations between OSTOX and diagnosis and AGE and sRAGE. Age, BMI, and TUD had no significant effects in this analysis. Tests for between-subject effects showed that there were significant associations between AGE and OSTOX and between sRAGE and diagnosis.

**Table 3.** Results of multivariate GLM analysis examining the associations between AGE and sRAGE, and oxidative stress toxicity (OSTOX) and diagnosis of healthy controls, and low methamphetamine (low-MA) and high-MA abuse

| Tests | Dependent variables | Explanatory variables | F | df | p |
| --- | --- | --- | --- | --- | --- |
| **Multivariate** | AGE; sRAGE | OSTOX | 5.328 | 2/85 | 0.007 |
|  |  | HC/low-MA/high-MA | 6.857 | 4/172 | <0.001 |
| **Between-subject effects** | AGE | OSTOX | 10.556 | 1/86 | 0.002 |
|  | sRAGE | OSTOX | 0.177 | 1/86 | 0.675 |
| **Between-subject effects** | AGE | HC/low-MA/high-MA | 0.477 | 2/86 | 0.622 |
|  | sRAGE | HC/low-MA/high-MA | 15.670 | 2/86 | <0.001 |

OSTOX: Index of oxidative stress, AGE: Advanced Glycation End Products, sRAGE: soluble receptor of advanced glycation end products,

### Intercorrelation matrix between the ARA, and biochemical and clinical data

The intercorrelation matrix between AGE, sRAGE, ARA and other biomarkers is shown in **Table 4**. AGE was significantly correlated with Castelli, AIP, OSTOX, OSTOX/ANTIOX, MAI symptom index, and FTD. sRAGE was significantly correlated with copper, Castelli, AIP, OSTOX, OSTOX/ANTIOX, MAI symptom index, psychosis, hostility, excitement, and FTD (all positively), magnesium and ANTIOX (inversely). ARA showed significant associations with copper, Castelli, AIP, OSTOX, OSTOX/ANTIOX, MAI symptom index, psychosis, hostility, excitement, FTD (all positively) and magnesium and ANTIOX (inversely). **Figure 1** shows the partial regression of the Castelli risk 1 index on ARA stress. Both Castelli (r=0.312, p=0.003, n=90) and AIP (r=0.316, p=0.002) were significantly correlated with OSTOX. Both atherogencity indices and ANTIOX contain HDLc and thus correlations between these variables are redundant.

**Table 4.**
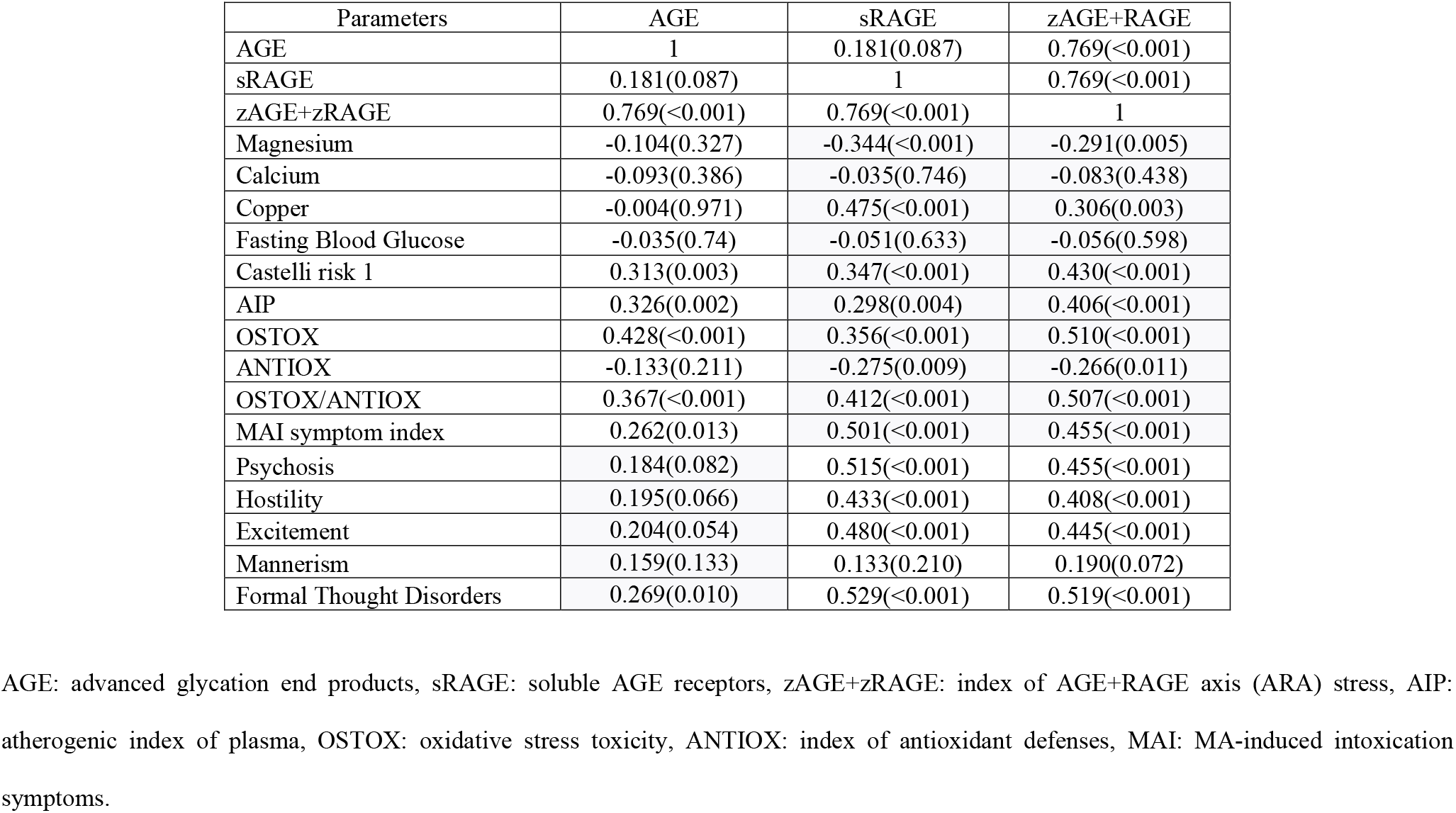
Intercorrelation matrix between AGE, sRAGE, ARA stress, biomarker and clical data in methamphetamine (MA) abuse and dependent patients and controls

| Parameters | AGE | sRAGE | zAGE+RAGE |
| --- | --- | --- | --- |
| AGE | 1 | 0.181(0.087) | 0.769(<0.001) |
| sRAGE | 0.181(0.087) | 1 | 0.769(<0.001) |
| zAGE+zRAGE | 0.769(<0.001) | 0.769(<0.001) | 1 |
| Magnesium | -0.104(0.327) | -0.344(<0.001) | -0.291(0.005) |
| Calcium | -0.093(0.386) | -0.035(0.746) | -0.083(0.438) |
| Copper | -0.004(0.971) | 0.475(<0.001) | 0.306(0.003) |
| Fasting Blood Glucose | -0.035(0.74) | -0.051(0.633) | -0.056(0.598) |
| Castelli risk 1 | 0.313(0.003) | 0.347(<0.001) | 0.430(<0.001) |
| AIP | 0.326(0.002) | 0.298(0.004) | 0.406(<0.001) |
| OSTOX | 0.428(<0.001) | 0.356(<0.001) | 0.510(<0.001) |
| ANTIOX | -0.133(0.211) | -0.275(0.009) | -0.266(0.011) |
| OSTOX/ANTIOX | 0.367(<0.001) | 0.412(<0.001) | 0.507(<0.001) |
| MAI symptom index | 0.262(0.013) | 0.501(<0.001) | 0.455(<0.001) |
| Psychosis | 0.184(0.082) | 0.515(<0.001) | 0.455(<0.001) |
| Hostility | 0.195(0.066) | 0.433(<0.001) | 0.408(<0.001) |
| Excitement | 0.204(0.054) | 0.480(<0.001) | 0.445(<0.001) |
| Mannerism | 0.159(0.133) | 0.133(0.210) | 0.190(0.072) |
| Formal Thought Disorders | 0.269(0.010) | 0.529(<0.001) | 0.519(<0.001) |

AGE: advanced glycation end products, sRAGE: soluble AGE receptors, zAGE+zRAGE: index of AGE+RAGE axis (ARA) stress, AIP: atherogenic index of plasma, OSTOX: oxidative stress toxicity, ANTIOX: index of antioxidant defenses, MAI: MA-induced intoxication symptoms.

**Figure 1.**
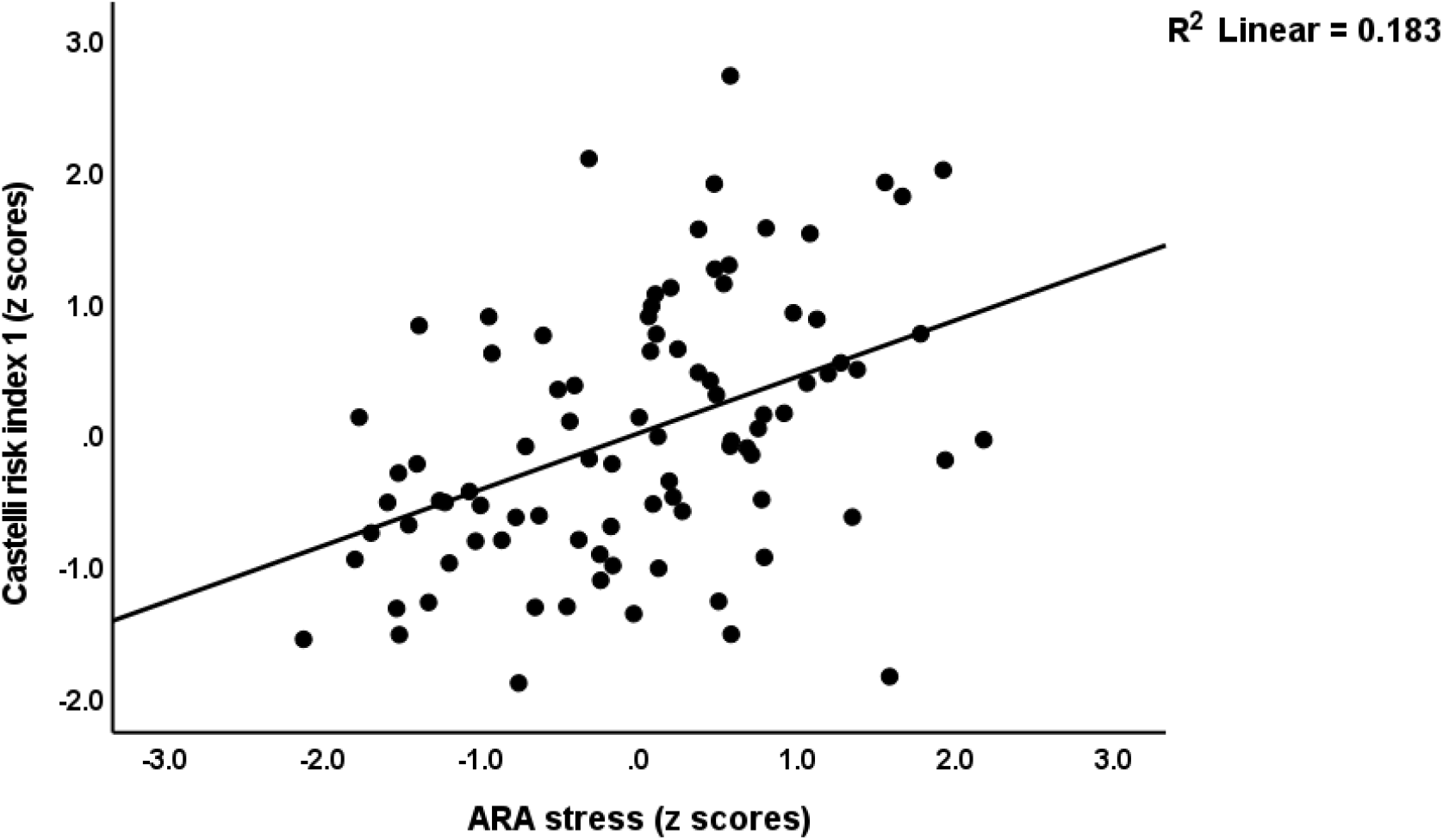
Partial regression of the Castelli risk index 1 on AGE-RAGE axis (ARA) stress.

**Electronic supplementary File (ESF) Table 1** shows the correlation of the psychiatric symptoms with the other serum biomarkers. The MAI symptom index was significantly correlated with copper, Castelli, AIP, and inversely with magnesium. All PHEM (psychosis, hostility, excitement, mannerism) symptom and FTD showed inverse associations with magnesium and positive with copper. In addition, psychosis showed a significant correlation with FBG, Castelli, and AIP. Hostility was significantly correlated with FBG, Castelli, and AIP (all positively). Excitement was significantly correlated with Castelli and AIP (all positively). Mannerism showed a significant correlation with FBG and Castelli, and FTD with Castelli and AIP.

### Results of the multiple regression analysis

**Table 5** presents the results of multiple regression analyses with neuro-psychiatric symptom domains as dependent variables and the measured biomarkers as explanatory variables. Regression #1 shows that 46.7% of the variance in the MAI symptom index was explained by the regression on ARA, copper, and TUD (all positively associated) and magnesium (negative association). **Figure 2** shows the partial regression of the MAI index on ARA. **Figure 3** shows the partial regression plot of the MAI symptom index on magnesium levels. Regression #2 shows that 51.1% of the variance in the psychosis score was explained by the regression on sRAGE, FBG, and TUD (all postively associated), and magnesium (negative association). Regression #3 shows that 41.7 % of the variance in hostility was explained by the regression on ARA and copper (both positively associated), and magnesium (negatively associated). Regression #4 shows that a considerable part of the variance in excitement (45.5%) was explained by the regression on sRAGE, FBG, TUD (all positively), and magnesium (inversely). Regression #5 shows that 12.8% of the variance in the mannerism was explained by FBG (positively associated) and magnesium (inversely associated). A significant part of the variance of FTD (52.3%) can be explained by the regression on ARA, copper, TUD, age (all positively associated), and magnesium (negatively associated).

**Table 5.** Results of multiple regression analyses with psychiatric symptom domains as dependent variables and the measured biomarkers as explanatory variables

| Dependent Variables | Explanatory Variables | Coefficients of input variables |  |  | Model statistics |  |  |  |
| --- | --- | --- | --- | --- | --- | --- | --- | --- |
| | | $\beta$ | t | p | R<sup>2</sup> | F | df | p |
| #1. MAI symptom index | **Model** |  |  |  | 0.467 | 18.62 | 4/85 | <0.001 |
|  | Magnesium | -0.273 | -2.95 | 0.004 |  |  |  |  |
|  | zAGE+zRAGE | 0.303 | 3.52 | <0.001 |  |  |  |  |
|  | Copper | 0.218 | 2.43 | 0.017 |  |  |  |  |
|  | TUD | 0.193 | 2.27 | 0.026 |  |  |  |  |
| #2. Psychosis | **Model** |  |  |  | 0.511 | 22.18 | 4/85 | <0.001 |
|  | Magnesium | -0.367 | -4.31 | <0.001 |  |  |  |  |
|  | sRAGE | 0.338 | 4.06 | <0.001 |  |  |  |  |
|  | FBG | 0.200 | 2.57 | 0.012 |  |  |  |  |
|  | TUD | 0.197 | 2.38 | 0.020 |  |  |  |  |
| #3. Hostility | **Model** |  |  |  | 0.417 | 20.49 | 3/86 | <0.001 |
|  | Magnesium | -0.419 | -4.52 | <0.001 |  |  |  |  |
|  | zAGE+zRAGE | 0.227 | 2.58 | 0.012 |  |  |  |  |
|  | Copper | 0.193 | 2.07 | 0.042 |  |  |  |  |
| #4. Excitement | **Model** |  |  |  | 0.455 | 17.75 | 4/85 | <0.001 |
|  | Magnesium | -0.342 | -3.80 | <0.001 |  |  |  |  |
|  | sRAGE | 0.305 | 3.47 | <0.001 |  |  |  |  |
|  | TUD | 0.214 | 2.45 | 0.016 |  |  |  |  |
|  | FBG | 0.180 | 2.19 | 0.031 |  |  |  |  |
| #5. Mannerism | **Model** |  |  |  | 0.128 | 6.39 | 2/87 | <0.001 |
|  | Magnesium | -0.261 | -2.58 | 0.012 |  |  |  |  |
|  | FBG | 0.207 | 2.05 | 0.044 |  |  |  |  |
| #6. Formal Thought Disorders | **Model** |  |  |  | 0.523 | 18.42 | 5/84 | <0.001 |
|  | Magnesium | -0.290 | -3.29 | 0.001 |  |  |  |  |
|  | zAGE+zRAGE | 0.325 | 3.97 | <0.001 |  |  |  |  |
|  | Copper | 0.247 | 2.88 | 0.005 |  |  |  |  |
|  | TUD | 0.169 | 2.09 | 0.040 |  |  |  |  |
|  | Age | 0.151 | 1.99 | 0.050 |  |  |  |  |
| #7. MAI symptom index | **Model** |  |  |  | 0.460 | 24.42 | 3/86 | <0.001 |
|  | OSTOX/ANTIOX | 0.317 | 3.42 | <0.001 |  |  |  |  |
|  | Magnesium | -0.284 | -3.16 | 0.002 |  |  |  |  |
|  | sRAGE | 0.273 | 3.08 | 0.003 |  |  |  |  |
| #8. MAI symptom index | **Model** |  |  |  | 0.475 | 25.93 | 3/86 | <0.001 |
|  | AROSTOX/ANTIOX | 0.425 | 4.77 | <0.001 |  |  |  |  |
|  | Magnesium | -0.261 | -2.88 | 0.005 |  |  |  |  |
|  | Copper | 0.179 | 1.99 | 0.049 |  |  |  |  |

AGE: advanced glycation end products, ANTIOX: index of antioxidant defenses, FBG: Fasting Blood Glucose, OSTOX: index of oxidative stress, MAI: MA intoxication symptoms, sRAGE: soluble receptor of advanced glycation end products, TUD: tobacco use disorder, AROSTOX/ANTIOX: AROSTOX/ANTIOX: index of zAGE+zRAGE and oxidative stress and lowered antioxidant defenses, zAGE+zRAGE: AGE-RAGE axis stress

**Figure 2.**
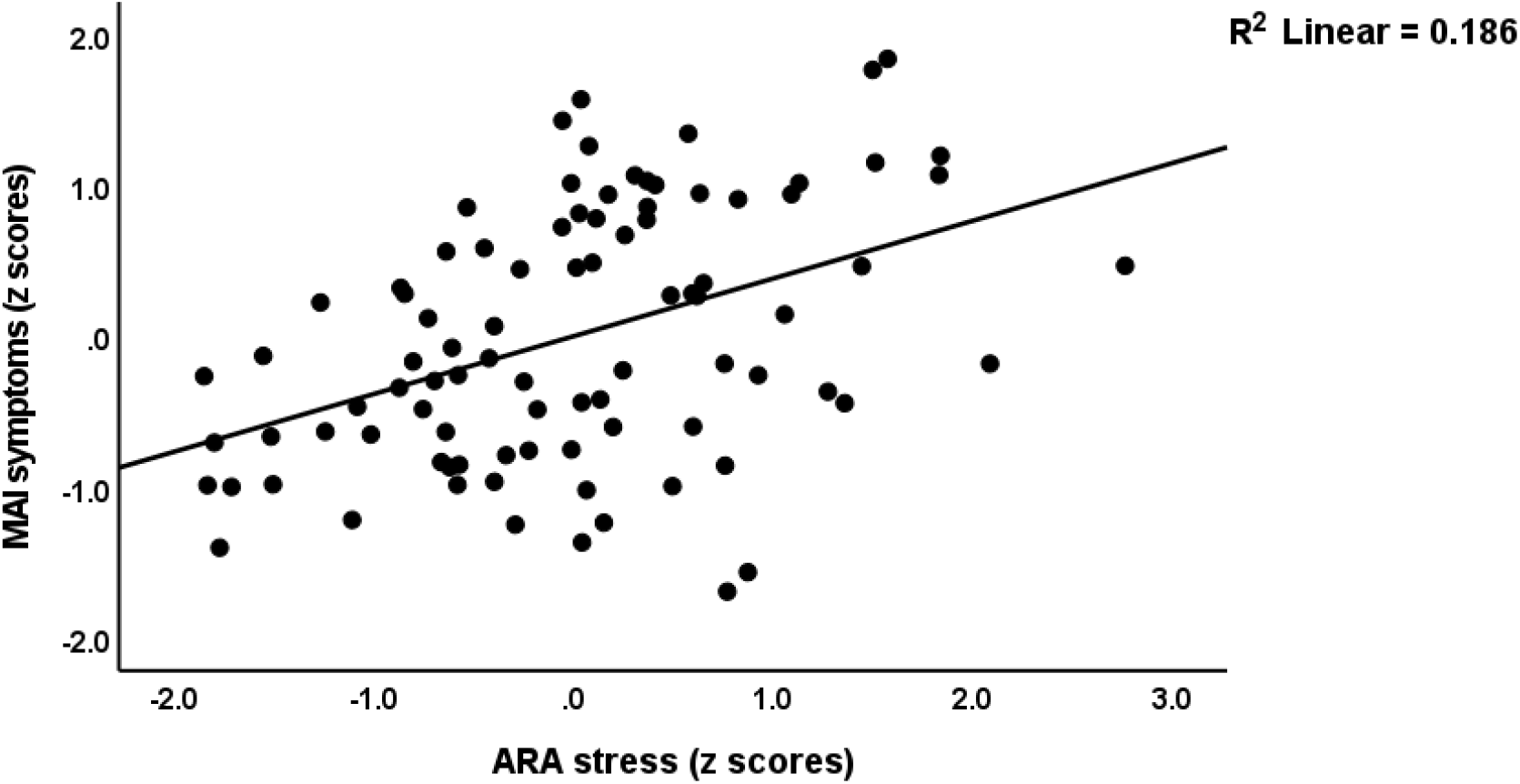
Partial regression of the methamphetamine intoxication (MAI) symtoms on AGE-RAGE axis (ARA) stress.

**Figure 3.**
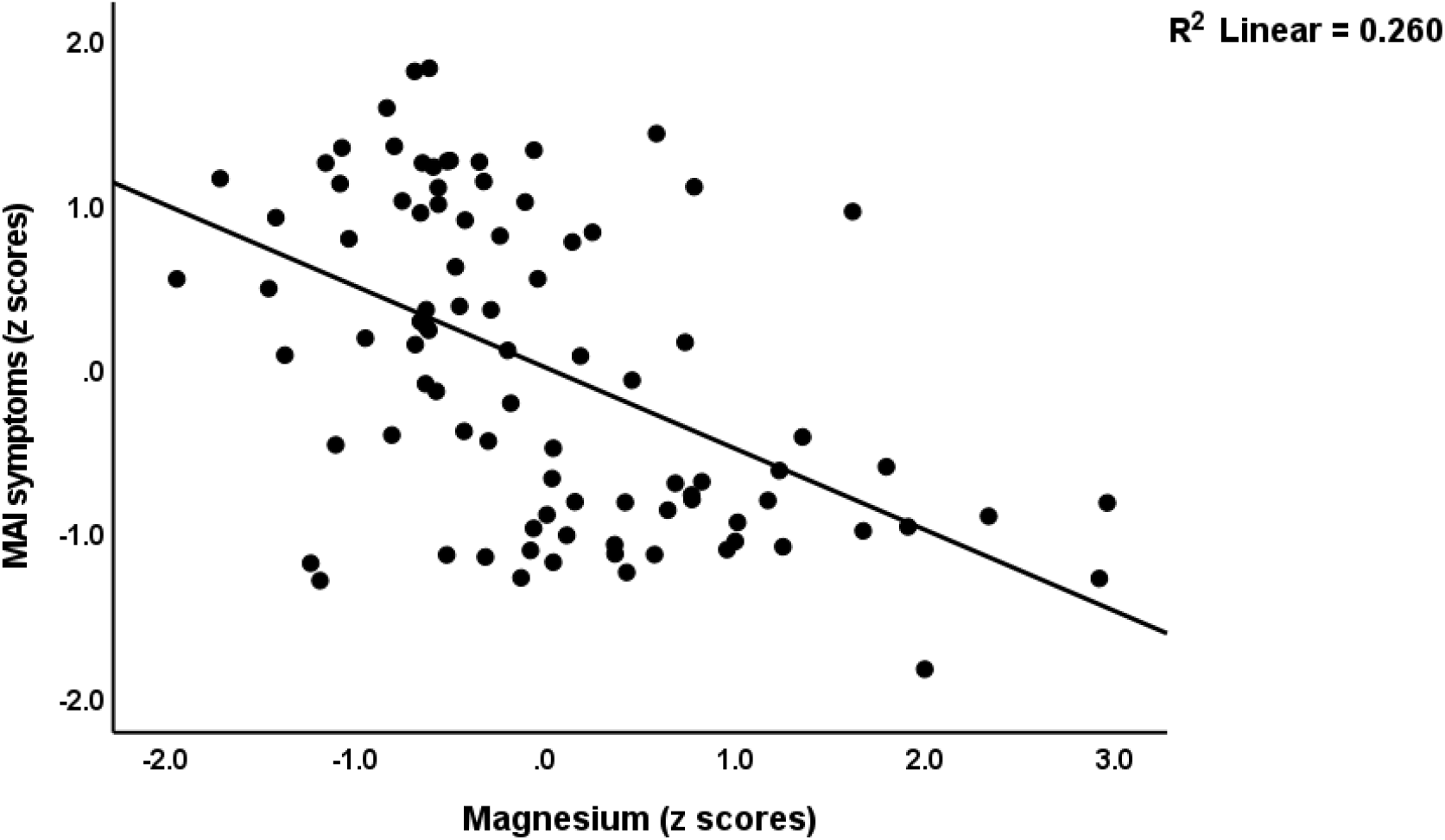
Partial regression of methamphetamine intoxication (MAI) symtoms on magnesium levels.

We have re-assessed the regressions with the MAI index as dependent variable and included the OSTOX/ANTIOX ratio. Regression #7 shows that a considerable part of the variance in the MAI index (46.0%) was explained by the regression on OSTOX/ANTIOX and sRAGE (both positively), and magnesium (inversely). Regression #8 shows that 47.5% of the variance in the MAI index was explained by the regression on AROSTOX/ANTIOX and copper (all showed positive associations) and magnesium that showed a negative association. **Figure 4** displays the partial regression of the MAI symptom index score on AROSTOX/ANTIOX.

**Figure 4.**
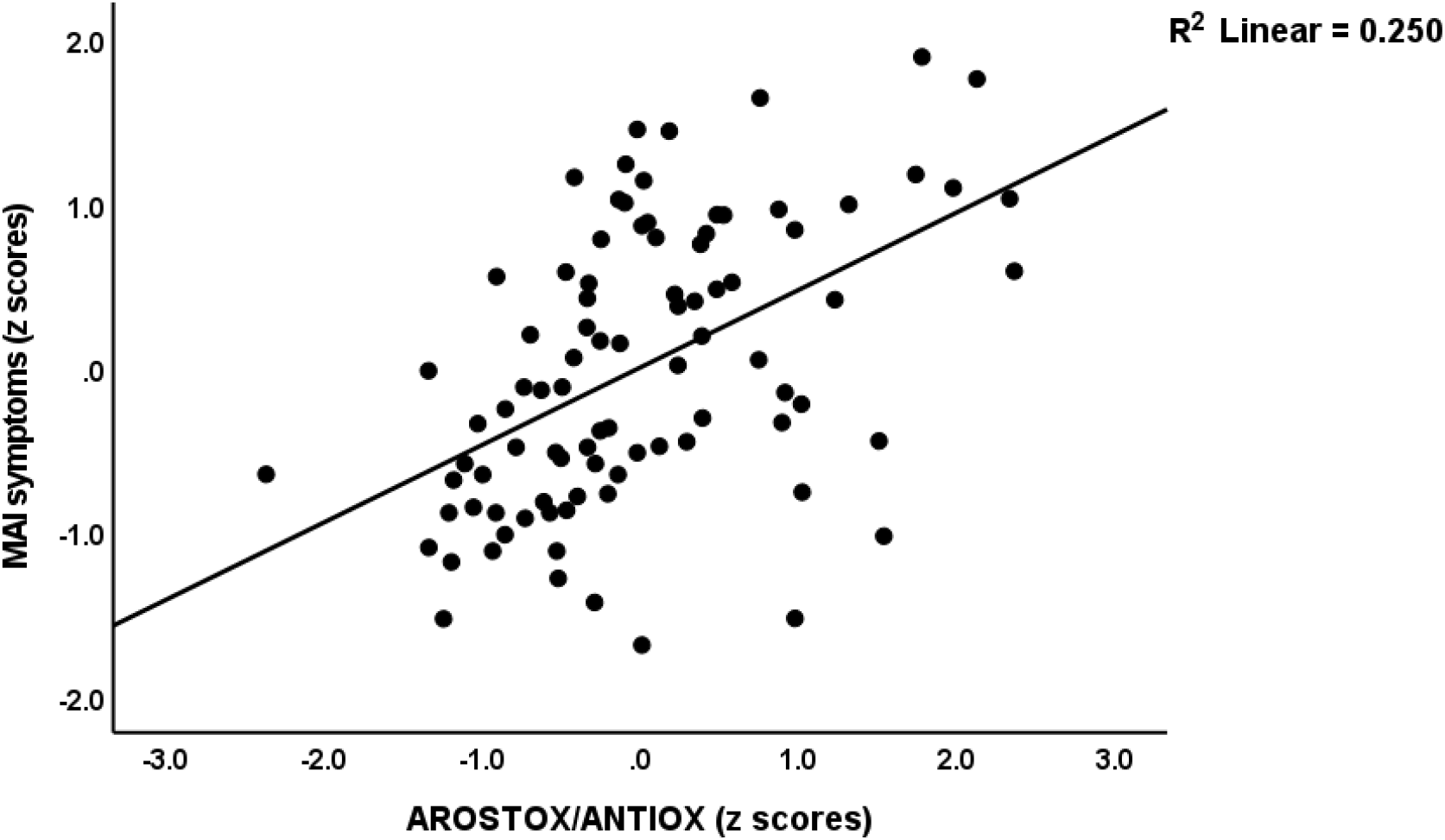
Partial regression of methamphetamine intoxication (MAI) symtoms on an integrated index of increased AGE-RAGE and oxidative stress and lowered antioxidant defenses (AROST/ANTIOX)

**Figure 5.**
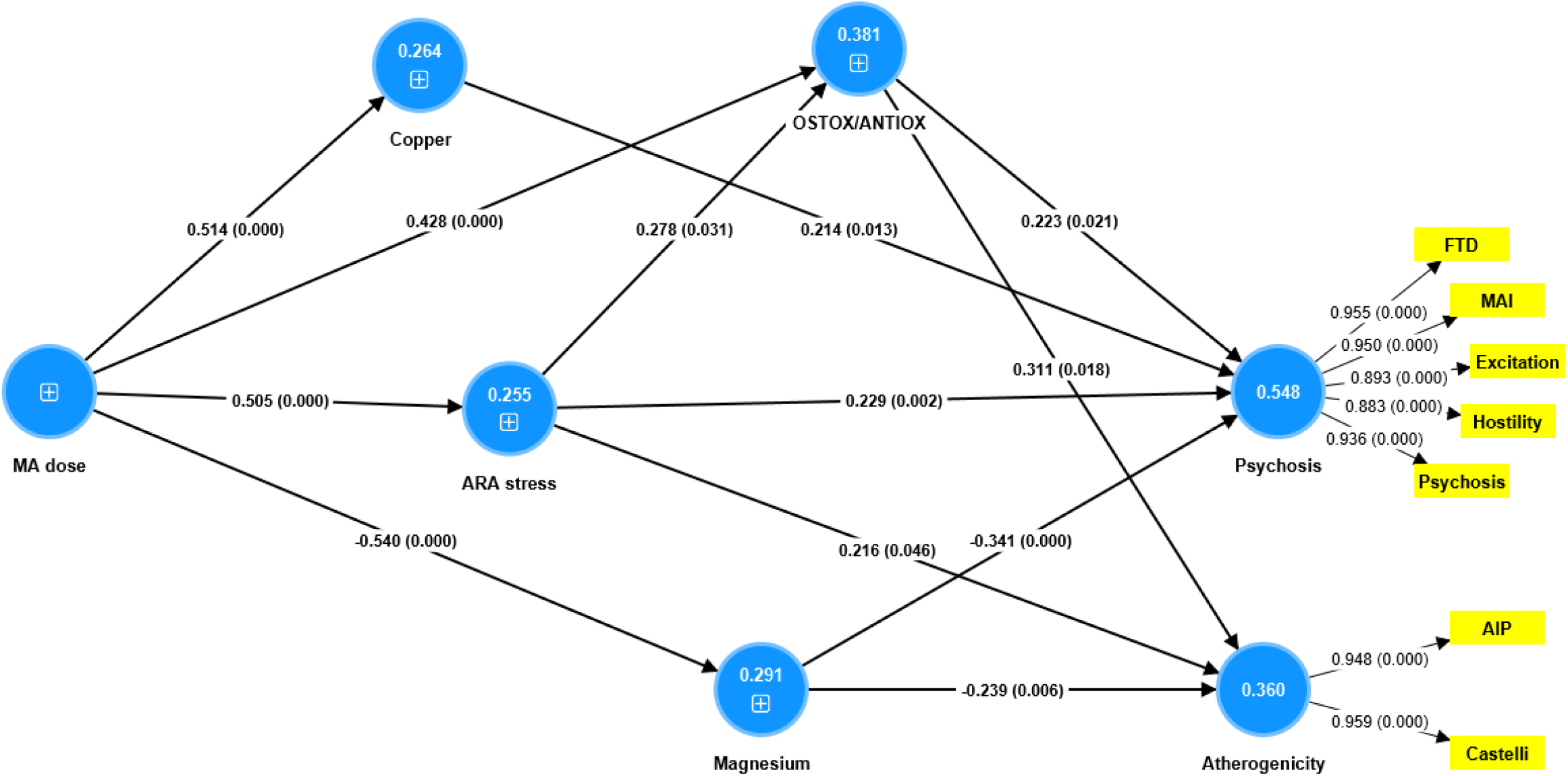
Results of Partial Least Squares analysis. A latent vector (LV) extracted from methamphetamine (MA)-induced psychotic symptoms is the final outcome, which is predicted by AGE-RAGE axis (ARA) stress, oxidative damage / antioxidant defenses (OSTOX/ANTIOX), copper and magnesium. The effects of MA dose on the symptoms are mediated by the 4 biomarkers. Three of the latter also predict atherogenicity, conceptualized as a latent vector extracted from the Castelli risk index 1 and the atherogenic index of plasma. Shown are path coefficients with exact p values, and loadings with p values. Figure in blue circles: explained variance. +: indicates single indicators.

### Correlation of MA SUD features with biomarkers

**Table 6** shows the intercorrelation matrix between PC_SDS and MA intake parameters and the measured biomarkers in MA SUD patients. PC_SDS was significantly correlated with sRAGE, copper, Castelli, AIP (all positively), and magnesium (inversely). The MA dose was significantly correlated with AGE, sRAGE, copper, Castelli, AIP (all positively), and magnesium (inversely). The MA route variable was significantly associated with AGE, sRAGE, copper, Castelli and AIP.

**Table 6.** Intercorrelation matrix between methamphetamine (MA) abuse parameters, including MA dose, MA route, and severity of MA dependence (PC_SDS) with biomarkers of MA use

| Parameters | PC_SDS | MA dose | MA Route |
| --- | --- | --- | --- |
| **MA dose** | 0.774 (<0.001) |  |  |
| **MA route** | 0.523 (<0.001) | 0.460 (<0.001) |  |
| **AGE** | 0.190 (0.073) | 0.215 (0.043) | 0.270 (0.010) |
| **sRAGE** | 0.519 (<0.001) | 0.632 (<0.001) | 0.389 (<0.001) |
| **zAGE+zRAGE** | 0.501 (<0.001) | 0.571 (<0.001) | 0.387 (<0.001) |
| **Magnesium** | -0.569 (<0.001) | -0.582 (<0.001) | -0.400 (<0.001) |
| **Calcium** | -0.093 (0.383) | -0.162 (0.130) | -0.213 (0.044) |
| **Copper** | 0.453 (<0.001) | 0.574 (<0.001) | 0.400 (<0.001) |
| **Fasting Blood Glucose** | 0.207 (0.050) | 0.190 (0.074) | 0.197 (0.063) |
| **Castelli** | 0.400 (<0.001) | 0.476 (<0.001) | 0.434 (<0.001) |
| **AIP** | 0.251 (0.017) | 0.416 (<0.001) | 0.307 (0.003) |

AGE: advanced glycation end products, AIP: atherogenic index of plasma, sRAGE: soluble receptor of advanced glycation end products, zAGE+zRAGE: index of AGE-RAGE axis stress

### Results of PLS analysis

Figure 6. shows the final PLS model after feature reduction. With SRMR = 0.027, the current PLS model was of adequate quality. The MIP symptom factor demonstrated adequate convergence and construct reliability with AVE=0.854, rho A=0.967, composite reliability=0.960, and Cronbach’s alpha=0.957, and all loadings were greater than 0.883 at p < 0.001. The atherogenicity LV model was adequate with AVE=0.909, rho A=0.909, composite reliability=0.967, and Cronbach’s alpha=0.957 while all loadings were > 0.948 at p<0.001. CTA verified that neither of the two latent vectors were incorrectly specified as reflective models. PLSPredict revealed that the construct indicator Q^2^ predict values were all greater than zero, indicating that the prediction error was less than that of the simplest benchmark. Complete PLS analysis using 5,000 bootstraps revealed that the regression of the MIP phenome on ARA, OSTOX/ANTIOX, magnesium and copper explained 54.8% of the variance. The atherogenicity index did not contribute to the phenome and 36.0% of its variance was explained by OSTOX/ANTIOX, ARA, and magnesium. MA dose explained a large part of the variances in ARA (25.5%), OSTOX/ANTIOX (38.1%), copper (26.4%) and magnesium (29.1%). There were significant total indirect effects of MA dose on MIP phenome (t=9.25, p<0.0001) that were mediated via ARA (t=2.38, p=0.018), magnesium (t=2.91, p=0.004), and copper (t=2.00, p=0.046). There were significant total indirect effects of MA dose on atherogenicity (t=8.48, p<0.0001) that were mediated via magnesium (t=2.49, p=0.013) and OSTOX/ANTIOX (t=2.13, p=0.033). Post-hoc power analysis performed on the primary outcome data (namely the phenome) showed that (given R^2^=0.548, n=90, 4 predictors, at alpha=0.05, two tailed) the obtained power was 1.

## Discussion

### Increased ARA stress in MA users and MIP

The first major finding of this study is that MA SUD is associated with elevated serum AGE and sRAGE concentrations, while MIP is characterized by elevated sRAGE levels and an elevated ARA index. In addition, an increase in sRAGE (but not AGE) and the ARA index is significantly correlated with the severity of MAI and PHEM symptoms and FTD. In addition, there was a significant correlation between the MA dose, MA dependence, and elevated sRAGE and AGE concentrations in patients with MA SUD. The sRAGE is shed from the cell surface receptor, and increased sRAGE concentrations in serum reflect an increase in cell surface RAGE receptor expression or overstimulation (44, 45). Serum AGE and sRAGE concentrations should be evaluated concurrently to assess the severity of RAGE signaling and ARA stress (31). In our study, the ARA score, therefore, reflects the severity of ARA stress.

It is well-established that MA has the potential to induce a psychotic state (46, 47), and both increased severity of MA abuse, dependence and dose are associated with an increased risk of MIP (11). The results of the current study indicate that MA SUD and dependence and dosage induce ARA stress, and that the latter contributes to the onset of MIP symptoms. A recent systematic review demonstrated an increase in serum AGE levels in schizophrenia patients (48). According to Arai et al. (49), schizophrenia is associated with a 1.7-fold increase in AGE levels and these results were confirmed by Miyahi et al. (50) and Kouidrat et al. (51). Intriguingly, a recent study shows that adolescents with low muscular strength are more likely to experience cognitive difficulties, which are most likely mediated by AGEs (52). According to Hagen et al. (2020), there is an association between significantly increased AGE accumulation and recent-onset psychosis (53). Katsuda et al. (54) and Emanue (55), on the other hand, were unable to detect an increase in AGE levels in schizophrenia or reported a decrease. After six weeks of treatment, Steiner et al. (56) observed an increase in sRAGE levels in schizophrenia patients, whereas Miyashita et al. (57) discovered that schizophrenia patients had lower sRAGE levels than controls. One study found a link between RAGE 429T/C polymorphisms and schizophrenia in Han Chinese (58). Psychoticism and schizophrenia are linked to the functional RAGE polymorphism Gly82Ser (rs2070600, 244G>A), which modulates increased sRAGE plasma levels (59).

There are multiple explanations for the role of ARA stress in psychosis pathophysiology. Due to its ability to bind oxytocin in the brain, RAGE is a crucial factor in social recognition (60). In addition, RAGE may cross the blood-brain barrier and may, therefore, modulate the release of oxytocin in the brain, which mediates prosocial behaviors when CD markers such as CD38 and CD157 are present (60). Second, the ARA axis is tightly linked to immune-inflammatory and oxidative and nitrosative stress (IO&NS) pathways, which play a crucial role in MIP (11) and the deficit phenotype of schizophrenia (61). In the following section, the associations between ARA stress and IO&NS pathways in MA and MIP are discussed.

### ARA stress and IO&NS pathways

The second major finding of his study is that ARA stress is strongly associated with oxidative stress indicators (OSTOX and OSTOX/ANTIOX) and with decreased ANTIOX defenses. Both AGE and sRAGE were significantly associated with OSTOX, but only sRAGE was associated with decreased ANTIOX defenses. The interaction of AGEs with their cell surface receptors may lead to an increase in pro-inflammatory cytokine production and oxidative stress via ROS production by NADPH oxidase in the mitochondria (62, 63, 64). RAGE-mediated intracellular signaling can lead, through activation of nuclear factor-κB (65), to increased expression of pro-inflammatory cytokines, including interleukin (IL)-1 and tumor necrosis factor (TNF)-, and RAGE itself (45, 66-68). Inflammatory signals such as HMGB1 (69), lipopolysaccharides, and TNF-α (70) induce RAGE shedding and the release of sRAGE.

However, circulating sRAGE may bind ligands such as AGEs, thereby offsetting the detrimental effects of AGE binding to their cell receptors (63). Not only is sRAGE a protective factor against inflammatory stress, but it also plays an antioxidant role by reducing ROS production (45, 71, 72) sRAGE may also act as a ligand of the leukocyte integrin MAC-1 and transduce pro-inflammatory signals, inducing leukocyte recruitment to injury or inflammation sites (73). Consequently, elevated sRAGE levels may be considered a potential biomarker of inflammation and oxidative stress and may reflect overstimulation of the ARA, whereas elevated sRAGE may exert anti-inflammatory and antioxidant effects ((45, 74).

Our results show that oxidative stress damage and ARA stress had independent and cumulative effects on psychotic symptoms, and that ARA stress also had significant indirect effects that were mediated by increased oxidative damage. Thus, it appears that not all the effects of the ARA axis on the onset of psychotic symptoms are due to O&NS pathways.

Other possibilities are increased levels of neurotoxic cytokines, including TNF-α, that were shown to play a role in deficit schizophrenia (75). ARA signaling and MA SUD are also involved in gut inflammation and increased gut permeability (leaky gut), which leads to an increase in the translocation of LPS, microbiota, and toxins into the peripheral circulation (76-78). Leaky gut may lead to activation of IO&NS pathways and blood-brain breakdown, thereby playing a crucial role in deficit schizophrenia (79). ARA stress also contributes to the production of RNS and subsequent nitrosative stress and peroxynitrite formation, which play a major role in MA-induced dopaminergic neurotoxicity, which can be mitigated by selective antioxidants and peroxynitrite decomposition catalysts (80, 81). Significantly, AGE production may stimulate ROS and RNS production, including peroxynitrite, resulting in a vicious cycle of nitrosative stress, oxidative protein damage, and diminished antioxidant defenses (26). Thus, the intertwined associations between MA-induced IO&NS and ARA stress play a crucial role in the development of psychotic symptoms and MIP.

In addition, the results of the current study indicate that MA SUD is associated with decreased magnesium levels and elevated copper levels, and that both magnesium and copper had significant independent effects on the severity of psychotic symptoms. In contrast, the current study did not reveal any effects of MA use on calcium levels, nor did it reveal any effects of calcium on psychotic symptoms or MIP. According to some reports, trace elements like copper, as well as ions like calcium and magnesium may play a role in schizophrenia. Thus, serum copper levels are elevated in patients with schizophrenia (82). Magnesium treatment may ameliorate psychosis in patients with schizophrenia who have decreased serum and intracellular magnesium levels (83). Furthermore, in patients with schizophrenia, magnesium deficiency is significantly associated with deficits in neurocognitive functions that predict more severe psychotic symptoms (36). Importantly, decreased zinc and magnesium are indicants of immune activation and an acute inflammatory response (38, 84).

According to our PLS analysis, the ARA axis, the OSTOX-ANTIOX ratio, elevated copper levels, and decreased magnesium levels could collectively account for up to 54.8% of the variance in psychotic symptoms. While 54.8% indicates a very large effect size, it also suggests that additional MA-related mechanisms may be involved in the onset of psychotic symptoms. For instance, MA may prevent the breakdown of excess dopamine and norepinephrine, leading to increased neurotoxicity via a variety of interdependent mechanisms, such as excitotoxicity and mitochondrial damage (85). In addition, MA has immunosuppressive properties that may fuel inflammatory responses, and MA may also cause microgliosis and blood-brain barrier breakdown (86-88). MA increases nuclear factor-κB in the brain via RAGE-mediated intracellular signalling that stimulates secretion of pro-inflammatory cytokines (e.g., IL1- and TNF) ((89, 90).

### ARA stress and atherogenicity

The third major finding of this study is that MA SUD is associated with increased atherogenicity, as indicated by an increased Castelli risk index 1 and AIP, and that increased atherogenicity in patients is strongly related to MA dependence severity and dose. In addition, both atherogenicity indices were strongly correlated with ARA stress, as well as oxidative damage. Moreover, both increased atherogenicity indices were significantly associated with the severity of psychotic symptoms, although this association was no longer significant after accounting for ARA stress, OSTOX, copper, and magnesium.

We discovered that the increased atherogenicity in MA SUD patients was a result of elevated total cholesterol, VLDL cholesterol, and triglyceride levels, whilst lower HDLc levels were a more specific indicator of MIP. In some studies, MA has been shown to increase serum cholesterol, LDLc, and triglycerides (91). In addition, there is a significant increase in serum triglycerides following MA administration in animal models, whilst, in cell lines, MA administration may induce cholesterol biosynthesis genes resulting in the accumulation of free cholesterol within the cell (91-93). Not all studies, however, reported such effects (94) (95).

The MA-induced changes in cholesterol homeostasis may in part explain increased atherosclerosis risk, atherosclerotic plaque formation, cardiovascular complications, and acute coronary syndromes (96) (92, 97) (98). Moreover, MA abuse may promote enhanced atherogenicity via different other mechanisms: a) increased catecholamine turnover, increased ROS and oxidative stress, enhanced proinflammatory gene expression and inflammatory toxicity, b) direct effects on cardiac and vascular tissues resulting in increased endothelial activation and plaque formation (98, 99), and c) mitochondrial dysfunctions (32, 100-103).

Importantly, the results of our PLS analysis indicate that increased atherogenicity may also be predicted by ARA stress and decreased magnesium levels. Both AGEs and sRAGE are considered biomarkers of atherosclerotic processes and cardiovascular disease, although elevated sRAGE may provide protection against cardiovascular disease (30, 31, 104, 105). Additionally, decreased magnesium and increased copper levels contribute independently to increased atherogenicity via distinct mechanisms (38).

## Conclusion

MA SUDs cause ARA stress, oxidative damage, and lipid peroxidation, as well as atherogenicity, lowered magnesium and increased copper. Increased sRAGE concentrations are strongly correlated with MA dependence severity and MA dose. Increased ARA stress is strongly correlated with increased lipid peroxidation and lowered antioxidant defenses and MA-induced intoxication symptoms, psychosis, hostility, excitement, and formal thought disorders. These biomarkers mediate the effects of increasing MA doses on MIP symptoms, explaining 54.8% of the variance. OSTOX/ANTIOX, ARA stress, and decreased magnesium explain 36.0% of the variance in the atherogenicity indices. MA SUD results in intertwined increases in ARA and oxidative stress, which predict the severity of MIP symptoms and increased atherogenicity. ARA and oxidative stress are drug targets to improve and prevent MIP and atherogenicity in patients with MA abuse and dependence.

## Data Availability

The dataset generated during and/or analyzed during the current study will be available from the corresponding author upon reasonable request and once the dataset has been fully exploited by the authors.

## Acknowledgment

We thank the staff of the Psychiatry Unit of Al-Hussein Medical City in the Kerbala Governorate for their help in the collection of samples. We also acknowledge the work of the high-skilled staff of Asia Clinical Laboratory in Najaf city for their help in the ELISA and spectroscopic measurements.

## Author’s contributions

All the contributing authors have participated in preparation of the manuscript.

## References

1. Ahmad FB CJ, Rossen LM, Sutton P. Provisional drug overdose death counts.. National Center for Health Statistics. 2022;Designed by LM Rossen, A Lipphardt, FB Ahmad, JM Keralis, and Y Chong: National Center for Health Statistics.

2. Faraone SV, Hess J, Wilens T. Prevalence and consequences of the nonmedical use of amphetamine among persons calling poison control centers. Journal of attention disorders. 2019;23(11):1219–28.

3. Jones CM, Houry D, Han B, Baldwin G, Vivolo-Kantor A, Compton WM. Methamphetamine use in the United States: epidemiological update and implications for prevention, treatment, and harm reduction. Annals of the New York Academy of Sciences. 2022;1508(1):3–22.

4. Hogarth S, Manning E, van den Buuse M. Chronic Methamphetamine and Psychosis Pathways. In: Patel VB, Preedy VR, editors. Handbook of Substance Misuse and Addictions: From Biology to Public Health. Cham: Springer International Publishing; 2021. p. 1–26.

5. Zhang Y, Lu C, Zhang J, Hu L, Song H, Li J, et al. Gender differences in abusers of amphetamine-type stimulants and ketamine in southwestern China. Addictive behaviors. 2013;38(1):1424–30.

6. Thomas E, Lategan H, Verster C, Kidd M, Weich L. Methamphetamine-induced psychosis: Clinical features, treatment modalities and outcomes. The South African journal of psychiatry : SAJP : the journal of the Society of Psychiatrists of South Africa. 2016;22(1):980.

7. Arunogiri S, Foulds JA, McKetin R, Lubman DI. A systematic review of risk factors for methamphetamine-associated psychosis. The Australian and New Zealand journal of psychiatry. 2018;52(6):514–29.

8. Yang M, Yang C, Liu T, London ED. Methamphetamine-associated psychosis: links to drug use characteristics and similarity to primary psychosis. International journal of psychiatry in clinical practice. 2020;24(1):31–7.

9. Grant KM, LeVan TD, Wells SM, Li M, Stoltenberg SF, Gendelman HE, et al. Methamphetamine-associated psychosis. Journal of Neuroimmune Pharmacology. 2012;7(1):113–39.

10. Arunogiri S, Foulds JA, McKetin R, Lubman DI. A systematic review of risk factors for methamphetamine-associated psychosis. Australian & New Zealand Journal of Psychiatry. 2018;52(6):514–29.

11. Al-Hakeim HK, Altufaili MF, Almulla AF, Moustafa SR, Maes M. Increased lipid peroxidation and lowered antioxidant defenses predict methamphetamine induced psychosis. Cells. 2022;11(22):3694.

12. Ikeda M, Okahisa Y, Aleksic B, Won M, Kondo N, Naruse N, et al. Evidence for shared genetic risk between methamphetamine-induced psychosis and schizophrenia. Neuropsychopharmacology. 2013;38(10):1864–70.

13. Gan H, Song Z, Xu P, Su H, Pan Y, Zhao M, et al. A comparison study of working memory deficits between patients with methamphetamine-associated psychosis and patients with schizophrenia. Shanghai Archives of Psychiatry. 2018;30(3):168.

14. Jayanthi S, Daiwile AP, Cadet JL. Neurotoxicity of methamphetamine: Main effects and mechanisms. Experimental Neurology. 2021;344:113795.

15. Lin M, Sambo D, Khoshbouei H. Methamphetamine regulation of firing activity of dopamine neurons. Journal of Neuroscience. 2016;36(40):10376–91.

16. Kim B, Yun J, Park B. Methamphetamine-induced neuronal damage: neurotoxicity and neuroinflammation. Biomolecules & therapeutics. 2020;28(5):381.

17. Hsieh JH, Stein DJ, Howells FM. The neurobiology of methamphetamine induced psychosis. Frontiers in Human Neuroscience. 2014;8.

18. Elkashef A, Vocci F, Hanson G, White J, Wickes W, Tiihonen J. Pharmacotherapy of methamphetamine addiction: an update. Substance Abuse. 2008;29(3):31–49.

19. Moszczynska A, Callan SP. Molecular, behavioral, and physiological consequences of methamphetamine neurotoxicity: implications for treatment. Journal of pharmacology and experimental therapeutics. 2017;362(3):474–88.

20. Limanaqi F, Gambardella S, Biagioni F, Busceti CL, Fornai F. Epigenetic effects induced by methamphetamine and methamphetamine-dependent oxidative stress. Oxidative medicine and cellular longevity. 2018;2018.

21. Afzali S, Fadaei F, Oftadeh A, Ranjbar A. Salivary Biomarkers of Oxidative Stress in Methamphetamine Users: A Case-Control Study. Novelty in Clinical Medicine. 2022;1(2):95–100.

22. Henning C, Glomb MA. Pathways of the Maillard reaction under physiological conditions. Glycoconjugate journal. 2016;33(4):499–512.

23. Uribarri J, Cai W, Peppa M, Goodman S, Ferrucci L, Striker G, et al. Circulating glycotoxins and dietary advanced glycation endproducts: two links to inflammatory response, oxidative stress, and aging. The Journals of Gerontology Series A: Biological Sciences and Medical Sciences. 2007;62(4):427–33.

24. Abate G, Marziano M, Rungratanawanich W, Memo M, Uberti D. Nutrition and AGE-ing: Focusing on Alzheimer’s Disease. Oxidative medicine and cellular longevity. 2017;2017.

25. Byun K, Yoo Y, Son M, Lee J, Jeong G-B, Park YM, et al. Advanced glycation end-products produced systemically and by macrophages: A common contributor to inflammation and degenerative diseases. Pharmacology & therapeutics. 2017;177:44–55.

26. Ott C, Jacobs K, Haucke E, Santos AN, Grune T, Simm A. Role of advanced glycation end products in cellular signaling. Redox biology. 2014;2:411–29.

27. Kierdorf K, Fritz G. RAGE regulation and signaling in inflammation and beyond. Journal of leukocyte biology. 2013;94(1):55–68.

28. San Martin A, Foncea R, Laurindo FR, Ebensperger R, Griendling KK, Leighton F. Nox1-based NADPH oxidase-derived superoxide is required for VSMC activation by advanced glycation end-products. Free Radical Biology and Medicine. 2007;42(11):1671–9.

29. Hudson BI, Lippman ME. Targeting RAGE signaling in inflammatory disease. Annual review of medicine. 2018;69:349–64.

30. Egaña-Gorroño L, López-Díez R, Yepuri G, Ramirez LS, Reverdatto S, Gugger PF, et al. Receptor for advanced glycation end products (RAGE) and mechanisms and therapeutic opportunities in diabetes and cardiovascular disease: insights from human subjects and animal models. Frontiers in cardiovascular medicine. 2020;7:37.

31. Prasad K. AGE-RAGE Stress and Coronary Artery Disease. Int J Angiol. 2021;30(1):4–14.

32. Kevil CG, Goeders NE, Woolard MD, Bhuiyan MS, Dominic P, Kolluru GK, et al. Methamphetamine use and cardiovascular disease: in search of answers. Arteriosclerosis, thrombosis, and vascular biology. 2019;39(9):1739–46.

33. Hossain KJ, Kamal MM, Ahsan M, Islam S. Serum antioxidant micromineral (Cu, Zn, Fe) status of drug dependent subjects: Influence of illicit drugs and lifestyle. Substance Abuse Treatment, Prevention, and Policy. 2007;2(1):1–7.

34. Nechifor M. Magnesium in addiction–a general view. Magnesium Research. 2018;31(3):90–8.

35. Abood MT, Almashhedy LA, editors. Elevated of Calcium and Sodium Levels as a Result of Methamphetamine Addiction, Causesand Consequence. IOP Conference Series: Materials Science and Engineering; 2020: IOP Publishing.

36. Al-Musawi AF, Al-Hakeim HK, Al-Khfaji ZA, Al-Haboby IH, Almulla AF, Stoyanov DS, et al. In schizophrenia, the effects of the IL-6/IL-23/Th17 axis on health-related quality of life and disabilities are partly mediated by generalized cognitive decline and the symptomatome. International Journal of Environmental Research and Public Health. 2022;19(22):15281.

37. Al-Hakeim HK, Alhusseini AF, Al-Dujaili A, Debnath M, Maes M. The interleukin-6/interleukin-23/Thelper-17-axis as a driver of neuro-immune toxicity in the major neurocognitive psychosis or deficit schizophrenia: a precision nomothetic psychiatry analysis. medRxiv. 2022.

38. Mousa RF, Smesam HN, Qazmooz HA, Al-Hakeim HK, Maes M. A pathway phenotype linking metabolic, immune, oxidative, and opioid pathways with comorbid depression, atherosclerosis, and unstable angina. CNS spectrums. 2022;27(6):676–90.

39. Edition F. Diagnostic and statistical manual of mental disorders. Am Psychiatric Assoc. 2013;21(21):591–643.

40. Friedewald WT, Levy RI, Fredrickson DS. Estimation of the concentration of low-density lipoprotein cholesterol in plasma, without use of the preparative ultracentrifuge. Clinical chemistry. 1972;18(6):499–502.

41. Gossop M, Darke S, Griffiths P, Hando J, Powis B, Hall W, et al. The Severity of Dependence Scale (SDS): psychometric properties of the SDS in English and Australian samples of heroin, cocaine and amphetamine users. Addiction (Abingdon, England). 1995;90(5):607–14.

42. Morelli NR, Maes M, Bonifacio KL, Vargas HO, Nunes SOV, Barbosa DS. Increased nitro-oxidative toxicity in association with metabolic syndrome, atherogenicity and insulin resistance in patients with affective disorders. Journal of Affective Disorders. 2021;294:410–9.

43. Maes M, Brinholi FF, Michelin A-P, Matsumoto A, Semeao L, Almulla AF, et al. Increased lipid peroxidation and lowered lipid-associated antioxidant defenses mediate the effects of the paraoxonase 1 (PON1) Q192R polymorphism on disabilities and final stroke core volume in mild and moderate stroke. medRxiv. 2022.

44. Koyama H, Yamamoto H, Nishizawa Y. RAGE and Soluble RAGE: Potential Therapeutic Targets for Cardiovascular Diseases. Molecular Medicine. 2007;13(11):625–35.

45. Erusalimsky JD. The use of the soluble receptor for advanced glycation-end products (sRAGE) as a potential biomarker of disease risk and adverse outcomes. Redox Biology. 2021;42:101958.

46. McKetin R, Lubman DI, Baker AL, Dawe S, Ali RL. Dose-related psychotic symptoms in chronic methamphetamine users: evidence from a prospective longitudinal study. JAMA psychiatry. 2013;70(3):319–24.

47. McKetin R. Methamphetamine psychosis: insights from the past. Addiction. 2018;113(8):1522–7.

48. Kouidrat Y, Amad A, Arai M, Miyashita M, Lalau J-D, Loas G, et al. Advanced glycation end products and schizophrenia: A systematic review. Journal of psychiatric research. 2015;66:112–7.

49. Arai M, Yuzawa H, Nohara I, Ohnishi T, Obata N, Iwayama Y, et al. Enhanced carbonyl stress in a subpopulation of schizophrenia. Archives of general psychiatry. 2010;67(6):589–97.

50. Miyashita M, Yamasaki S, Ando S, Suzuki K, Toriumi K, Horiuchi Y, et al. Fingertip advanced glycation end products and psychotic symptoms among adolescents. npj Schizophrenia. 2021;7(1):1–6.

51. Kouidrat Y, Amad A, Desailloud R, Diouf M, Fertout E, Scoury D, et al. Increased advanced glycation end-products (AGEs) assessed by skin autofluorescence in schizophrenia. Journal of psychiatric research. 2013;47(8):1044–8.

52. Suzuki A, Yabu A, Nakamura H. Advanced glycation end products in musculoskeletal system and disorders. Methods. 2022;203:179–86.

53. Hagen JM, Sutterland AL, Edrisy S, Tan HL, de Haan L. Accumulation rate of advanced glycation end products in recent onset psychosis: a longitudinal study. Psychiatry Research. 2020;291:113192.

54. Katsuta N, Ohnuma T, Maeshima H, Takebayashi Y, Higa M, Takeda M, et al. Significance of measurements of peripheral carbonyl stress markers in a cross-sectional and longitudinal study in patients with acute-stage schizophrenia. Schizophrenia bulletin. 2014;40(6):1366–73.

55. Emanuele E, Martinelli V, Carlin MV, Fugazza E, Barale F, Politi P. Serum levels of soluble receptor for advanced glycation endproducts (sRAGE) in patients with different psychiatric disorders. Neuroscience letters. 2011;487(1):99–102.

56. Steiner J, Walter M, Wunderlich MT, Bernstein H-G, Panteli B, Brauner M, et al. A new pathophysiological aspect of S100B in schizophrenia: potential regulation of S100B by its scavenger soluble RAGE. Biological psychiatry. 2009;65(12):1107–10.

57. Miyashita M, Watanabe T, Ichikawa T, Toriumi K, Horiuchi Y, Kobori A, et al. The regulation of soluble receptor for AGEs contributes to carbonyl stress in schizophrenia. Biochemical and biophysical research communications. 2016;479(3):447–52.

58. Fu J, Zuo X, Yin J, Luo X, Li Z, Lin J, et al. Association of polymorphisms of the receptor for advanced glycation endproducts gene with schizophrenia in a Han Chinese population. BioMed Research International. 2017;2017.

59. Suchankova P, Klang J, Cavanna C, Holm G, Nilsson S, Jönsson EG, et al. Is the Gly82Ser polymorphism in the RAGE gene relevant to schizophrenia and the personality trait psychoticism? Journal of Psychiatry and Neuroscience. 2012;37(2):122–8.

60. Higashida H, Hashii M, Tanaka Y, Matsukawa S, Higuchi Y, Gabata R, et al. CD38, CD157, and RAGE as molecular determinants for social behavior. Cells. 2019;9(1):62.

61. Maes M, Kanchanatawan B. In (deficit) schizophrenia, a general cognitive decline partly mediates the effects of neuro-immune and neuro-oxidative toxicity on the symptomatome and quality of life. CNS spectrums. 2022;27(4):506–15.

62. Vincent AM, Perrone L, Sullivan KA, Backus C, Sastry AM, Lastoskie C, et al. Receptor for advanced glycation end products activation injures primary sensory neurons via oxidative stress. Endocrinology. 2007;148(2):548–58.

63. Prasad K. Low levels of serum soluble receptors for advanced glycation end products, biomarkers for disease state: myth or reality. Int J Angiol. 2014;23(1):11–6.

64. Cepas V, Collino M, Mayo JC, Sainz RM. Redox signaling and advanced glycation endproducts (AGEs) in diet-related diseases. Antioxidants. 2020;9(2):142.

65. C Tobon-Velasco J, Cuevas E, A Torres-Ramos M. Receptor for AGEs (RAGE) as mediator of NF-kB pathway activation in neuroinflammation and oxidative stress. CNS & Neurological Disorders-Drug Targets (Formerly Current Drug Targets-CNS & Neurological Disorders). 2014;13(9):1615–26.

66. Luan Z-G, Zhang H, Yang P-T, Ma X-C, Zhang C, Guo R-X. HMGB1 activates nuclear factor-κB signaling by RAGE and increases the production of TNF-α in human umbilical vein endothelial cells. Immunobiology. 2010;215(12):956–62.

67. Luan Z-G, Zhang H, Ma X-C, Zhang C, Guo R-X. Role of high-mobility group box 1 protein in the pathogenesis of intestinal barrier injury in rats with severe acute pancreatitis. Pancreas. 2010;39(2):216–23.

68. Holms RD. Long COVID (PASC) Is Maintained by a Self-Sustaining Pro-Inflammatory TLR4/RAGE-Loop of S100A8/A9> TLR4/RAGE Signalling, Inducing Chronic Expression of IL-1b, IL-6 and TNFa: Anti-Inflammatory Ezrin Peptides as Potential Therapy. Immuno. 2022;2(3):512–33.

69. Raucci A, Cugusi S, Antonelli A, Barabino SM, Monti L, Bierhaus A, et al. A soluble form of the receptor for advanced glycation endproducts (RAGE) is produced by proteolytic cleavage of the membrane-bound form by the sheddase a disintegrin and metalloprotease 10 (ADAM10). The FASEB Journal. 2008;22(10):3716–27.

70. Miyoshi A, Koyama S, Sasagawa-Monden M, Kadoya M, Konishi K, Shoji T, et al. JNK and ATF4 as two important platforms for tumor necrosis factor-α–stimulated shedding of receptor for advanced glycation end products. The FASEB Journal. 2019;33(3):3575–89.

71. Liu Y, Yu M, Zhang L, Cao Q, Song Y, Liu Y, et al. Soluble receptor for advanced glycation end products mitigates vascular dysfunction in spontaneously hypertensive rats. Molecular and cellular biochemistry. 2016;419(1-2):165–76.

72. Wu F, Feng JZ, Qiu YH, Yu FB, Zhang JZ, Zhou W, et al. Activation of receptor for advanced glycation end products contributes to aortic remodeling and endothelial dysfunction in sinoaortic denervated rats. Atherosclerosis. 2013;229(2):287–94.

73. Wang Y, Wang H, Piper MG, McMaken S, Mo X, Opalek J, et al. sRAGE induces human monocyte survival and differentiation. The Journal of Immunology. 2010;185(3):1822–35.

74. Dozio E, Vettoretti S, Lungarella G, Messa P, Corsi Romanelli MM. Sarcopenia in chronic kidney disease: Focus on advanced glycation end products as mediators and markers of oxidative stress. Biomedicines. 2021;9(4):405.

75. Maes M, Sirivichayakul S, Matsumoto AK, Maes A, Michelin AP, de Oliveira Semeão L, et al. Increased Levels of Plasma Tumor Necrosis Factor-α Mediate Schizophrenia Symptom Dimensions and Neurocognitive Impairments and Are Inversely Associated with Natural IgM Directed to Malondialdehyde and Paraoxonase 1 Activity. Mol Neurobiol. 2020;57(5):2333–45.

76. Prakash MD, Tangalakis K, Antonipillai J, Stojanovska L, Nurgali K, Apostolopoulos V. Methamphetamine: effects on the brain, gut and immune system. Pharmacological research. 2017;120:60–7.

77. Persons AL, Bradaric BD, Dodiya HB, Ohene-Nyako M, Forsyth CB, Keshavarzian A, et al. Colon dysregulation in methamphetamine self-administering HIV-1 transgenic rats. PLoS One. 2018;13(1):e0190078.

78. Snelson M, Lucut E, Coughlan MT. The Role of AGE-RAGE Signalling as a Modulator of Gut Permeability in Diabetes. Int J Mol Sci. 2022;23(3).

79. Maes M, Vojdani A, Sirivichayakul S, Barbosa DS, Kanchanatawan B. Inflammatory and oxidative pathways are new drug targets in multiple episode schizophrenia and leaky gut, Klebsiella pneumoniae, and C1q immune complexes are additional drug targets in first episode schizophrenia. Molecular neurobiology. 2021;58(7):3319–34.

80. Imam SZ, EL-YAZAL J, Newport GD, Itzhak Y, Cadet JL, Slikker Jr W, et al. Methamphetamine-induced dopaminergic neurotoxicity: role of peroxynitrite and neuroprotective role of antioxidants and peroxynitrite decomposition catalysts. Annals of the New York Academy of Sciences. 2001;939(1):366–80.

81. Riddle EL, Fleckenstein AE, Hanson GR. Mechanisms of methamphetamine-induced dopaminergic neurotoxicity. The AAPS journal. 2006;8:E413–E8.

82. Wolf TL, Kotun J, Meador-Woodruff JH. Plasma copper, iron, ceruloplasmin and ferroxidase activity in schizophrenia. Schizophrenia research. 2006;86(1-3):167–71.

83. Ordak M, Bulska E, Jablonka-Salach K, Luciuk A, Maj-Żurawska M, Matsumoto H, et al. Effect of disturbances of zinc and copper on the physical and mental health status of patients with alcohol dependence. Biological trace element research. 2018;183(1):9–15.

84. Maes M, Vandoolaeghe E, Neels H, Demedts P, Wauters A, Meltzer HY, et al. Lower serum zinc in major depression is a sensitive marker of treatment resistance and of the immune/inflammatory response in that illness. Biol Psychiatry. 1997;42(5):349–58.

85. Halpin LE, Collins SA, Yamamoto BK. Neurotoxicity of methamphetamine and 3,4-methylenedioxymethamphetamine. Life sciences. 2014;97(1):37–44.

86. Miyazaki I, Asanuma M, Diaz-Corrales FJ, Miyoshi K, Ogawa N. Direct evidence for expression of dopamine receptors in astrocytes from basal ganglia. Brain research. 2004;1029(1):120–3.

87. Martins T, Baptista S, Gonçalves J, Leal E, Milhazes N, Borges F, et al. Methamphetamine transiently increases the blood–brain barrier permeability in the hippocampus: role of tight junction proteins and matrix metalloproteinase-9. Brain research. 2011;1411:28–40.

88. Frausto RF, Chung DD, Boere PM, Swamy VS, Duong HN, Kao L, et al. ZEB1 insufficiency causes corneal endothelial cell state transition and altered cellular processing. PLoS One. 2019;14(6):e0218279.

89. Nakajima A, Yamada K, Nagai T, Uchiyama T, Miyamoto Y, Mamiya T, et al. Role of tumor necrosis factor-α in methamphetamine-induced drug dependence and neurotoxicity. Journal of Neuroscience. 2004;24(9):2212–25.

90. Coelho-Santos V, Leitão RA, Cardoso FL, Palmela I, Rito M, Barbosa M, et al. The TNF-α/Nf-κ B Signaling Pathway has a Key Role in Methamphetamine–Induced Blood–Brain Barrier Dysfunction. Journal of Cerebral Blood Flow & Metabolism. 2015;35(8):1260–71.

91. Koriem KM, Soliman RE. Chlorogenic and caftaric acids in liver toxicity and oxidative stress induced by methamphetamine. Journal of toxicology. 2014;2014.

92. Funakoshi T, Aki T, Tajiri M, Unuma K, Uemura K. Necroptosis-like Neuronal Cell Death Caused by Cellular Cholesterol Accumulation. The Journal of biological chemistry. 2016;291(48):25050–65.

93. Aoki S, Funakoshi T, Aki T, Uemura K. Aggregation-prone A53T mutant of α-synuclein exaggerates methamphetamine neurotoxicity in SH-SY5Y cells: Protective role of cellular cholesterol. Toxicology reports. 2022;9:2020–9.

94. Suriyaprom K, Tanateerabunjong R, Tungtrongchitr A, Tungtrongchitr R. Alterations in malondialdehyde levels and laboratory parameters among methamphetamine abusers. Journal of the Medical Association of Thailand= Chotmaihet thangphaet. 2011;94(12):1533–9.

95. Zhang M, Lv D, Zhou W, Ji L, Zhou B, Chen H, et al. The levels of triglyceride and total cholesterol in methamphetamine dependence. Medicine. 2017;96(16).

96. Rao LN, Ponnusamy T, Philip S, Mukhopadhyay R, Kakkar VV, Mundkur L. Hypercholesterolemia induced immune response and inflammation on progression of atherosclerosis in Apob tm2Sgy Ldlr tm1Her/J mice. Lipids. 2015;50(8):785–97.

97. Hawley LA, Auten JD, Matteucci MJ, Decker L, Hurst N, Beer W, et al. Cardiac complications of adult methamphetamine exposures. The Journal of emergency medicine. 2013;45(6):821–7.

98. Gao B, Li L, Zhu P, Zhang M, Hou L, Sun Y, et al. Chronic administration of methamphetamine promotes atherosclerosis formation in ApoE-/-knockout mice fed normal diet. Atherosclerosis. 2015;243(1):268–77.

99. Zhu P, Li L, Gao B, Zhang M, Wang Y, Gu Y, et al. Impact of chronic methamphetamine treatment on the atherosclerosis formation in ApoE-/-mice fed a high cholesterol diet. Oncotarget. 2017;8(33):55064–72.

100. Bauch H-J, Grünwald J, Vischer P, Gerlach U, Hauss W. A possible role of catecholamines in atherogenesis and subsequent complications of atherosclerosis. Experimental pathology. 1987;31(4):193–204.

101. Rezvan A, Ni CW, Alberts-Grill N, Jo H. Animal, in vitro, and ex vivo models of flow-dependent atherosclerosis: role of oxidative stress. Antioxidants & redox signaling. 2011;15(5):1433–48.

102. Lee YW, Hennig B, Yao J, Toborek M. Methamphetamine induces AP-1 and NF-κB binding and transactivation in human brain endothelial cells. Journal of neuroscience research. 2001;66(4):583–91.

103. Ramirez SH, Potula R, Fan S, Eidem T, Papugani A, Reichenbach N, et al. Methamphetamine disrupts blood–brain barrier function by induction of oxidative stress in brain endothelial cells. Journal of Cerebral Blood Flow & Metabolism. 2009;29(12):1933–45.

104. Melin EO, Dereke J, Hillman M. Higher levels of the soluble receptor for advanced glycation end products and lower levels of the extracellular newly identified receptor for advanced glycation end products were associated with lipid-lowering drugs in patients with type 1 diabetes: a comparative cross-sectional study. Lipids in Health and Disease. 2020;19(1):1–10.

105. Pinto RS, Minanni CA, de Araújo Lira AL, Passarelli M. Advanced Glycation End Products: A Sweet Flavor That Embitters Cardiovascular Disease. International Journal of Molecular Sciences. 2022;23(5):2404.

